# Cross-sectional and Longitudinal Relationship between Sex Hormones and Six Epigenetic Clocks in Older Adults: Results of the Berlin Aging Study II (BASE-II)

**DOI:** 10.1101/2024.11.15.24317371

**Authors:** Hannah Schmid, Valentin Max Vetter, Jan Homann, Vivien Bahr, Christina M. Lill, Vera Regitz-Zagrosek, Lars Bertram, Ilja Demuth

**Author notes:** **Corresponding author:** Ilja Demuth (Ph.D.) Charité - Universitätsmedizin Berlin Lipid Clinic at the Interdisciplinary Metabolism Center, Biology of Aging Group Augustenburger Platz 1 13353 Berlin.

## Abstract

**Background:** Beyond their essential roles in regulating reproduction and development, sex hormones play a crucial role in the aging processes. Observational studies have indicated that low sex hormone concentrations in older age are associated with adverse health events. DNA methylation age acceleration (DNAmAA) estimated from epigenetic clocks quantifies differences in biological aging. DNAmAA was previously shown to be associated with age at menopause, ovariectomy, hormone replacement therapy and testosterone level.

**Methods:** We analysed the relationship between estradiol, dehydroepiandrosterone sulfate (DHEAS) and the Free Androgen Index (FAI) with DNAmAA estimators from six epigenetic clocks (Horvath’s, Hannum’s, 7-CpG clock, PhenoAge, GrimAge, DunedinPACE) in 1,404 participants of the Berlin Aging Study II (BASE-II, mean age at baseline 68.7 ±3.7 years, 48% women). The relationship was investigated in multiple linear regression models cross-sectionally at two time points and longitudinally over on average 7.3 years of follow-up.

**Results:** We did not observe any consistent associations between the sex hormones and DNAmAA estimators investigated. However, we found several nominal associations (alpha=0.05) of unclear relevance. For instance, we identified an inverse association between DHEAS and Horvath’s DNAmAA, i.e. a reduced biological age with higher DHEAS levels in men at baseline. In women we found an inverse association between estradiol and DunedinPACE (baseline) and a positive association with GrimAge (follow-up). In longitudinal analyses, ΔDHEAS and ΔDunedinPACE were inversely associated in both sexes.

**Conclusions:** Our results suggest that sex hormones play at best a minor role with respect to biological aging in the older population studied here.

## Introduction

Biological aging refers to differences in the aging process in people of the same chronological age. One of the latest and most promising biomarker of aging is the DNA methylation age (DNAmA) and its deviation from chronological age, DNAmA acceleration (DNAmAA) ^1,2^, which are calculated from DNA methylation data by epigenetic clock algorithms.

To date, numerous epigenetic clocks are available that mostly differ in the way they were developed and the number and location of cytosine-phosphate-guanine (CpG) sites they include ^3-8^. Previous analyses showed differences in the clock’s ability to predict morbidity and age-related pathologies independent of chronological age ^1,9,10^. First generation clocks such as Horvath’s, Hannum’s and the 7-CpG clock are designed to predict chronological age ^3-5^. Second generation clocks like GrimAge and PhenoAge are highly correlated with age associated pathologies and lifespan ^6,7^. DunedinPACE, a third generation clock, is designed to capture an individual’s pace of aging as well as differences from a normal aging rate ^8^.

Sex hormones have been recognised as factors involved in the aging processes, in addition to their essential function in regulating reproduction and development ^11,12^. In general, estrogens such as estradiol are distinguished from gestagens and androgens including testosterone and dehydroepiandrosterone sulfate (DHEAS). Over the life course, humans experience profound changes in sex hormone concentrations ^12^. In women, estradiol declines rapidly to approximately 20% of its maximum premenopausal concentration after the menopause ^13^. In contrast, the testosterone concentration in men peaks around the age of thirty followed by a gradual decrease of one to two percent per year ^14^. In both sexes, the DHEAS level decreases slowly starting in the third decade and resulting in a reduction by approximately 80% of its maximum around the seventh decade ^15^. The close temporal association between the decline in sex hormones and the age-related loss of physical abilities suggests a causal interaction ^12^. This assumption is further supported by findings from observational studies that indicate an association of low sex hormone concentrations in older age with adverse health events ^16-18^. Some studies suggest that the decline in DHEAS is associated with a higher mortality risk in men ^19,20^. DHEAS was referred to as an “anti-aging hormone” ^21^ and is available as an over-the-counter supplement in some countries although there is no consensus on its effectiveness ^22^. Low levels of testosterone were suggested as a risk factor for short-and long-term mortality in older men ^23-25^. In older women, a long reproductive period and late onset of menopause are related to longevity and reduced risk of mortality ^26-28^. Premature loss of ovarian estrogens is associated with multimorbidity, while estrogen substitution showed protective effects ^29^.

Because of these properties and associations, the relationship of sex hormones and biological age is of high interest. Since effects of sex hormones on overall DNAm levels were shown before, an association of sex hormone concentrations and DNAmAA seems biologically plausible ^30,31^. Indeed, studies in mice showed an accelerated epigenetic age after ovariectomy^32^ and in postmenopausal women, associations between early age at menopause and higher GrimAge and Horvath’s DNAmAA have been reported ^7,33^. Furthermore, Levine and colleagues suggested that biological aging processes accelerate when women enter the post-reproductive phase and that postmenopausal use of hormone replacement therapy (HRT) decreases epigenetic age in some tissues ^33^. A recent study found an inverse association between GrimAge and PhenoAge DNAmAA, respectively, and testosterone in men ^34^. Since stronger associations were reported for bioavailable testosterone ^34^, we used the Free Androgen Index (FAI) as an indicator of the free, SHBG-unbound proportion of testosterone in our study ^35^.

Despite these initial findings suggesting an association between sex hormones and DNAmAA, their potential cross-sectional and longitudinal relationship at different time points has not been studied yet. Thus, the objective of this study was to analyse the relationship between sex hormones (estradiol, DHEAS and the FAI) with DNAmAA estimators employing six different epigenetic clocks at two time points about seven years apart and to investigate the change of these variables over the observation period. Based on the previously reported results discussed above, we hypothesised that sex hormone concentrations are inversely associated with DNAmAA and that the association is also reflected in our longitudinal data.

## Methods

### Study population

Participants of the Berlin Aging Study II (BASE-II) cohort were recruited from the greater metropolitan area of Berlin, Germany. The multidisciplinary BASE-II aims to identify mechanisms and factors contributing to a healthy aging process. A subsample consisting of 1,671 older participants (aged ≥60 years) from the BASE-II were medically assessed at baseline between 2009 and 2014 ^36^. As part of the GendAge study, 1083 participants were reassessed with a mean follow-up time of 7.4 years (SD=1.5) between 2018 and 2020 ^37^. Cohort characteristics are shown in Table 1 and Supplementary Table 1.

**Table 1:** Characteristics of the 1,404 BASE-II participants at baseline (T0) of which 1,015 were reassessed on average 7.3 years later (T1) as included in the current analyses.

| Variables | Men |  |  | Women |  |  |
| --- | --- | --- | --- | --- | --- | --- |
|  | Mean | SD | n | Mean | SD | n |
|  | Mdn | IQR |  | Mdn | IQR |  |
| Age T0 (years) | 69.02 | 3.78 | 731 | 68.41 | 3.58 | 673 |
| Age T1 (years) | 75.50 | 4.01 | 514 | 75.66 | 3.55 | 501 |
| Interval T0-T1 (years) | 7.02 | 1.37 | 514 | 7.63 | 1.46 | 501 |
| Pack years T0<sup>a</sup> | 5.00 | 21.25 | 702 | 0.00 | 6.69 | 652 |
| Alcohol T0 (g/d)<sup>a</sup> | 12.73 | 21.30 | 717 | 5.32 | 10.65 | 656 |
| BMI T0 (kg/m<sup>2</sup>) | 27.25 | 3.61 | 727 | 26.54 | 4.69 | 668 |
| Sex hormone level T0<sup>a</sup> |  |  |  |  |  |  |
| Estradiol (pmol/l) | 84.45 | 53.49 | 725 | 9.20 | 33.89 | 667 |
| DHEAS (ng/ml) | 1055.50 | 819.60 | 728 | 736.25 | 739.84 | 667 |
| FAI | 36.69 | 13.38 | 718 | 0.90 | 1.25 | 641 |
| DNAmA T0 (years) |  |  |  |  |  |  |
| Horvath | 65.17 | 4.97 | 488 | 62.81 | 4.64 | 468 |
| Hannum | 56.54 | 4.36 | 488 | 53.89 | 4.28 | 468 |
| 7-CpG clock | 67.64 | 7.68 | 714 | 65.02 | 7.39 | 635 |
| PhenoAge | 54.92 | 5.43 | 488 | 52.53 | 5.48 | 468 |
| GrimAge | 72.17 | 3.92 | 488 | 68.55 | 3.84 | 468 |

Note. <sup>a</sup>=Median + Inter Quartile Range, SD=Standard Deviation, Mdn=Median, IQR= Inter Quartile Range, FAI=Free Androgen Index, DNAmA=DNA methylation age (years).

### Sample selection

Composition of participant numbers and final analytical datasets at baseline and follow-up are represented in Figure 1. The follow-up data set comprises 17 additional participants who were not medically examined in the baseline assessment. Women who reported taking oral or transdermal estrogen-containing HRT within one year prior to the baseline assessment were excluded from the baseline and longitudinal analyses. At follow-up no information on HRT cessation was available. Following the maximum duration of HRT recommended in the literature, we did not exclude women from the follow-up analyses ^38^. DHEAS data of three men and one woman were excluded from the baseline dataset due to extremely high values (> 7 SD from the mean).

**Figure 1:**
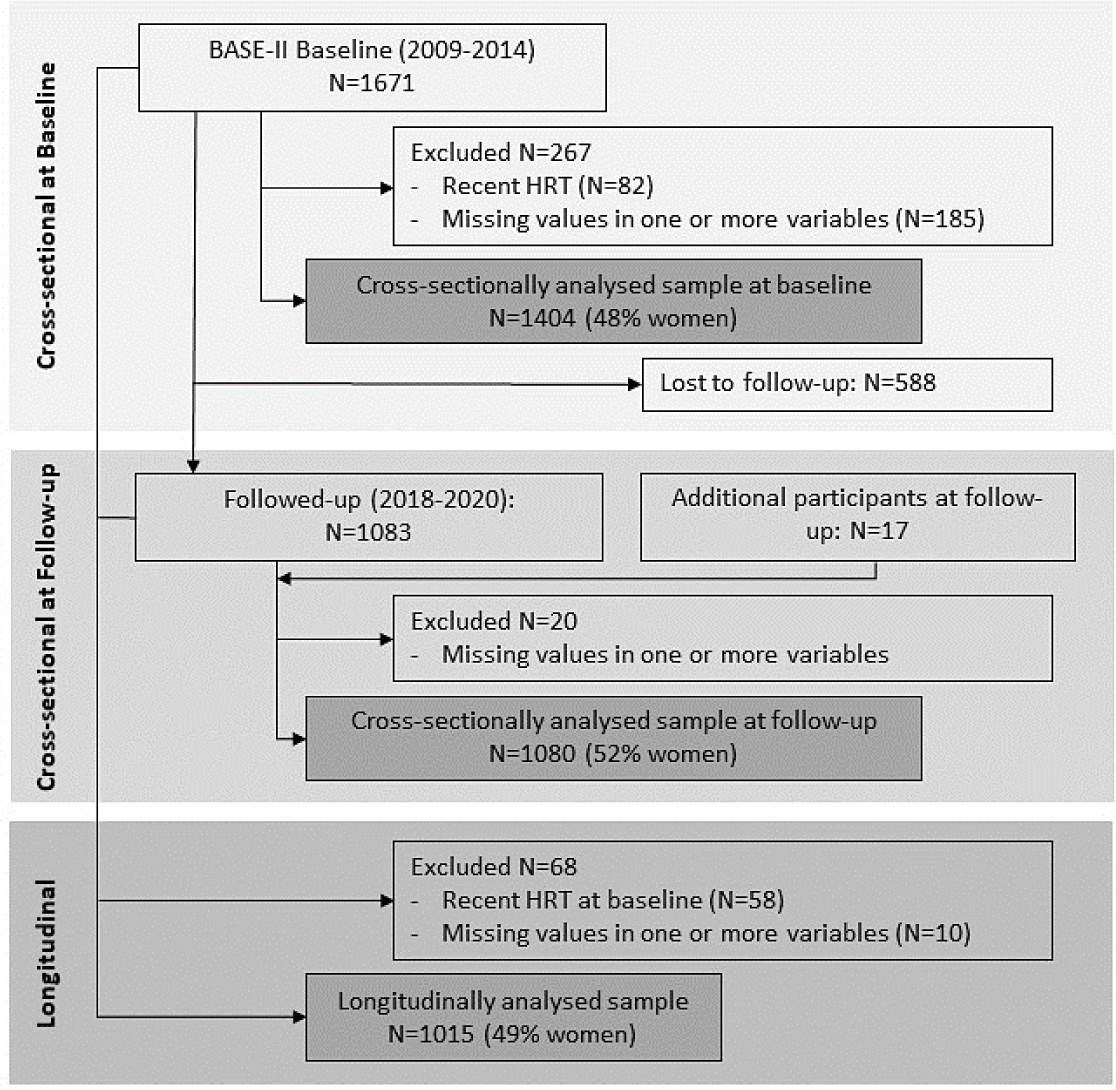
Case selection and final analytic datasets. Selected participants of the Berlin Aging Study II (BASE-II) were reassessed on average after 7.3 years. If the number of participants varied due to differences in the availability of hormone or epigenetic clock data, the maximum number of observations was reported in this flow chart. Individual sample sizes are indicated with each statistical analysis.

### DNA Methylation Data

Estimation of DNAmA was based on three first-generation (Horvath’s ^3^, Hannum’s ^4^, 7-CpG clock ^5^), two second-generation (PhenoAge ^6^, GrimAge ^7^) and one third-generation epigenetic clock (DunedinPACE ^8^). DNAm data originated from leukocyte DNA from whole blood samples drawn at baseline and follow-up assessment. Blood samples for sex hormone measurement were taken from the same blood draws. Methylation data was obtained using two methods. For Horvath’s ^3^, Hannum’s ^4^, PhenoAge ^6^, GrimAge ^7^ methylation age and DunedinPACE ^8^, methylation data was measured with the “Infinium MethylationEPIC” array (Illumina, Inc., USA). Detailed information about the sample handling and data processing were reported before ^39^. DunedinPACE was calculated as instructed in the original publication ^8^. A more detailed description of the methods can be found in ref. ^39^. For the 7-CpG clock measurement ^40^, DNAm data was obtained using methylation-sensitive single nucleotide primer extension (MS-SNuPE). For a detailed description of the laboratory protocol see reference ^5,40^. DNAmAA was calculated as the unstandardized residual resulting from regressing calculated DNAmA on chronological age while adjusting for blood cell counts (neutrophils, monocytes, lymphocytes, eosinophils) ^9,40^. Blood cell composition was measured by an accredited laboratory (Labor Berlin GmbH, Berlin, Germany) using flow cytometry.

### Sex hormones

Blood was drawn from all participants after an >8h fasting period and kept at 4–8 °C until analysis on the same day. Sex hormones were measured in serum samples by an accredited laboratory (Labor Berlin GmbH, Berlin, Germany) using fluoro-(estradiol, total testosterone (TT)), radio-(DHEAS) and chemiluminescentimmunoassays (sex hormone-binding globulin (SHBG)). During the study period, laboratory methods for measurement of sex hormones changed to electrochemiluminescence immunoassays (ECLIA). For further details we refer to the Supplementary Methods. In the final dataset, n=152 (estradiol, TT and SHBG) and n=39 (DHEAS) participants were affected by the change. A comparison of methods conducted by the accredited laboratory showed a high degree of comparability between the methods used (R^2^=0.95 and 0.97, data not shown). The FAI was calculated by using the formula: TT (nmol/l) × 100 / SHBG (nmol/l) ^35^.

### Covariates

Covariates were selected based on commonly employed factors within the field ^34^. Smoking behaviour was assessed in pack years during a one-on-one interview by trained study personnel. Daily intake of alcohol (g/d) was assessed using the European Prospective Investigation into Cancer and Nutrition Food Frequency Questionnaire (EPIC-FFQ) ^41^. Body Mass Index (BMI, kg/m2) was calculated from height and weight measured with an electronic measuring station (seca 763 measuring station, SECA, Germany). Information on menopausal status, medication and use of HRT, including initiation and cessation of HRT, were self-reported and based on medical reports when available.

### Statistical analysis

Statistical analyses were carried out using the IBM SPSS software package (IBM Corp. Released 2023. IBM SPSS Statistics for Windows, Version 29.0.2.0 Armonk, NY: IBM Corp). To account for sex-dependent hormone profiles, all statistical procedures were carried out in sex-stratified subgroups. Distribution of sex hormone data was visually inspected and found to be skewed. Therefore, non-parametric tests were conducted. Spearman correlation coefficients were calculated. The DNAmAA estimators were treated as a dependent variable in the multiple linear regression analyses. Linear regression models were adjusted for chronological age, BMI, pack years and alcohol intake (g/d). For longitudinal analyses, change in DNAmAA and sex hormones (denoted by Δvariable) was calculated as the difference between values at follow-up and baseline examination and found to be normally distributed. Pearson correlation coefficients were calculated. Linear regression analyses of ΔDNAmAA on Δsex hormones were adjusted for baseline variables (chronological age, sex hormone concentration, DNAmAA, pack years, alcohol intake (g/d), BMI and years since baseline assessment). Only participants with no missing values in the variables included in the respective analyses were considered (available case analysis). This resulted in varying numbers of complete cases depending on the clock and hormone analysed. A p-value below 0.05 was considered as statistically significant. Due to the explorative character of the study we did not correct for multiple testing.

## Results

### Descriptive Statistics

The baseline sample consisted of 1,404 participants (48% women). As part of the follow-up examination 1,015 participants (49% women) were reassessed on average 7.3 years (SD=1.5) later. The mean age at baseline was 68.4 years (SD=3.6) for women and 69.0 years (SD=3.8) for men. At baseline all women were postmenopausal with a mean interval of 18.7 years since menopause (SD=7.3, n=668) and 49.7 (SD=6.0) years as the average age at menopause. In men and women, median DHEAS concentration and the FAI decreased significantly between the assessments (Wilcoxon signed-rank test, p<0.001, Table 2). An increase in median estradiol concentration was found among men (p<0.001). A statistically significant increase in GrimAge DNAmAA by 0.38 years was shown for men (paired t-test p<0.001, Table 2), while GrimAge DNAmAA decreased significantly by 0.33 years in women (p=0.004). In both sexes, there was a significant increase in DunedinPACE by 0.07 years (p<0.001).

**Table 2:** Sex hormone level and DNAmAA estimators of participants who attended both baseline and follow-up assessments.

|  | Men |  |  |  | Women |  |  |  |
| --- | --- | --- | --- | --- | --- | --- | --- | --- |
|  | T0 (2009-2014) | T1 (2018-2020) |  |  | T0 (2009-2014) | T1 (2018-2020) |  |  |
| Variables | Mean (SD) | Mean (SD) | n | p-value | Mean (SD) | Mean (SD) | n | p-value |
|  | Mdn (IQR) | Mdn (IQR) |  |  | Mdn (IQR) | Mdn (IQR) |  |  |
| **DNAmAA (years)** |  |  |  |  |  |  |  |  |
| Horvath | 0.62 (4.21) | 0.46 (3.98) | 443 | 0.258 | -0.61 (4.07) | -0.54 (3.69) | 400 | 0.651<sup>b</sup> |
| Hannum | 0.71 (3.46) | 0.81 (4.00) | 443 | 0.414 | -0.77 (3.31) | -0.74 (3.53) | 400 | 0.794<sup>b</sup> |
| 7-CpG clock | 1.05 (6.86) | 0.97 (5.49) | 449 | 0.735 | -1.22 (6.66) | -0.96 (5.30) | 402 | 0.275<sup>b</sup> |
| PhenoAge | 0.31 (4.48) | 0.67 (5.42) | 443 | 0.065 | -0.29 (4.54) | -0.61 (5.04) | 400 | 0.104<sup>b</sup> |
| GrimAge | 1.11 (3.00) | 1.50 (3.23) | 443 | <0.001 | -1.02 (2.73) | -1.34 (2.86) | 400 | 0.004<sup>b</sup> |
| DunedinPACE (biol./chron.) | 1.03 (0.11) | 1.10 (0.12) | 490 | <0.001 | 0.98 (0.10) | 1.05 (0.11) | 468 | <0.001<sup>b</sup> |
| **Sex hormone level<sup>a</sup>** |  |  |  |  |  |  |  |  |
| Estradiol (pmol/l) | 84.00 (53.78) | 97.40 (48.30) | 495 | <0.001 | 9.20 (29.77) | 9.20 (25.10) | 487 | 0.786<sup>c</sup> |
| DHEAS (ng/ml) | 1074.28 (847.38) | 889.00 (699.0) | 499 | <0.001 | 743.20 (682.27) | 643.00 (604.80) | 486 | <0.001<sup>c</sup> |
| FAI | 37.08 (13.99) | 26.32 (9.91) | 491 | <0.001 | 0.89 (1.19) | 0.50 (0.82) | 466 | <0.001<sup>c</sup> |

Note. <sup>a</sup>=Median (Inter Quartile Range), <sup>b</sup>=paired t-test, <sup>c</sup>=Wilcoxon signed-rank test, Mdn=Median, IQR=Inter Quartile Range, SD=Standard Deviation, DNAmAA=DNA methylation age acceleration (years), FAI=Free Androgen Index, T0=Baseline assessment, T1=Follow-up assessment. A Wilcoxon signed-rank test was used to test for differences in median sex hormone concentration between baseline and follow-up assessment. To test for differences in mean DNAmAA a paired t-test was calculated. Only participants with available data at both time points are included in this table.

### Cross-sectional analyses: baseline

Baseline analyses indicated weak correlations (Spearman’s rho) between sex hormones and DNAmAA estimators ranging from r=-0.06 to 0.13 for men and from r=-0.07 to 0.10 for women (Supplementary Table 2). After adjustment for potential confounders an inverse association of DHEAS with Horvath’s DNAmAA (β=-0.0007, p=0.032, Table 3) was found in linear regression analyses for men. To illustrate, a decrease in Horvath’s DNAmAA by 0.45 years for every increase in DHEAS of one SD was observed. In women, estradiol was inversely associated with DunedinPACE (β=-0.0004, p=0.032, Table 3). Every one SD increase in estradiol was associated with a 0.01 year decrease in DunedinPACE. Cross-sectional analyses on the baseline dataset revealed no further relevant associations (see Supplementary Table 3).

**Table 3:** Multiple linear regression analyses of DNAmAA on estradiol, DHEAS and FAI for older participants of the BASE-II at baseline.

| Hormone | DNAmAA | Model | Estimate | SE | Men |  | p-value | n | Estimate | SE | Women |  | p-value | n |
| --- | --- | --- | --- | --- | --- | --- | --- | --- | --- | --- | --- | --- | --- | --- |
|  |  |  |  |  | CI<br>LL | CI<br>UL |  |  |  |  | CI<br>LL | CI<br>UL |  |  |
| Estradiol | Horvath | 1 | 0.007 | 0.005 | -0.002 | 0.015 | 0.141 | 446 | 0.008 | 0.007 | -0.006 | 0.022 | 0.288 | 406 |
|  |  | 2 | 0.004 | 0.005 | -0.005 | 0.013 | 0.390 | 420 | 0.004 | 0.007 | -0.010 | 0.019 | 0.571 | 384 |
|  | GrimAge | 1 | 0.001 | 0.003 | -0.005 | 0.007 | 0.741 | 446 | 0.002 | 0.005 | -0.008 | 0.011 | 0.740 | 406 |
|  |  | 2 | 0.002 | 0.003 | -0.003 | 0.008 | 0.400 | 420 | -0.004 | 0.005 | -0.013 | 0.005 | 0.414 | 384 |
|  | DunedinPACE | 1 | 2.7E-5 | 1.1E-4 | -1.9E-4 | 2.4E-4 | 0.802 | 483 | -1.6E-4 | 1.7E-4 | -4.9E-4 | 1.7E-4 | 0.346 | 464 |
|  |  | 2 | 6.6E-6 | 1.1E-4 | -2.1E-4 | 2.2E-4 | 0.952 | 455 | -3.6E-4 | 1.7E-4 | -0.001 | -3.0E-5 | 0.032 | 433 |
| DHEAS | Horvath | 1 | -4.8E-4 | 3.3E-4 | -0.001 | 1.6E-4 | 0.144 | 449 | -3.1E-4 | 4.0E-4 | -0.001 | 5.E-4 | 0.441 | 405 |
|  |  | 2 | -0.001 | 3.4E-4 | -0.001 | -6.2E-5 | 0.032 | 424 | -4.3E-4 | 4.2E-4 | -0.001 | 4.E-4 | 0.300 | 383 |
|  | GrimAge | 1 | 1.2E-4 | 2.3E-4 | -3.3E-4 | 0.001 | 0.603 | 449 | 3.2E-4 | 2.6E-4 | -2.E-4 | 0.001 | 0.219 | 405 |
|  |  | 2 | -9.0E-5 | 2.1E-4 | -0.001 | 3.3E-4 | 0.677 | 424 | 2.9E-4 | 2.5E-4 | -2.1E-4 | 0.001 | 0.260 | 383 |
|  | DunedinPACE | 1 | -1.8E-6 | 7.8E-6 | -1.7E-5 | 1.3E-5 | 0.817 | 486 | 4.5E-6 | 9.5E-6 | -1.E-5 | 2.E-5 | 0.637 | 464 |
|  |  | 2 | -8.6E-7 | 7.8E-6 | -1.6E-5 | 1.4E-5 | 0.912 | 459 | 1.2E-5 | 9.3E-6 | -6.E-6 | 3.E-5 | 0.185 | 433 |
| FAI | Horvath | 1 | 0.011 | 0.016 | -0.021 | 0.043 | 0.503 | 444 | -0.020 | 0.141 | -0.297 | 2.6E-1 | 0.888 | 387 |
|  |  | 2 | 0.004 | 0.017 | -0.029 | 0.038 | 0.799 | 418 | -0.046 | 0.144 | -0.328 | 0.237 | 0.751 | 367 |
|  | GrimAge | 1 | -0.026 | 0.011 | -0.049 | -0.004 | 0.021 | 444 | 0.019 | 0.094 | -0.166 | 0.203 | 0.842 | 387 |
|  |  | 2 | -0.019 | 0.011 | -0.041 | 0.002 | 0.072 | 418 | 0.028 | 0.088 | -0.145 | 0.202 | 0.748 | 367 |
|  | DunedinPACE | 1 | -0.001 | 4.E-4 | -0.001 | 2.E-4 | 0.123 | 480 | 0.003 | 0.003 | -0.004 | 0.009 | 0.382 | 443 |
|  |  | 2 | -2.E-4 | 4.E-4 | -0.001 | 0.001 | 0.613 | 452 | 0.001 | 0.003 | -0.005 | 0.007 | 0.737 | 415 |

Model 1: unadjusted; Model 2: adjusted for chronological age, smoking (pack years), alcohol (g/d), BMI. Note: DNAmAA=DNA methylation age
acceleration, SE=Standard Error, CI=Confidence Interval, LL=Lower Limit, UL=Upper Limit.

### Cross-sectional analyses: follow-up

Similar to baseline analyses, weak correlations were found between follow-up DNAmAA estimators and sex hormone level ranging from r=-0.11 to 0.10 for men and r=-0.08 to 0.09 for women (Supplementary Table 2). Adjusted linear regression analyses for women revealed a weak positive association between estradiol and the GrimAge DNAmAA (β=0.002, p=0.036, Supplementary Table 4). All other cross-sectional analyses on the follow-up dataset did not show any recurrent associations between sex hormones and DNAmAA (see Supplementary Table 4).

### Longitudinal analyses

To assess whether changes in sex hormone concentrations were related to changes in DNAmAA over the follow-up period, we conducted longitudinal analyses. Weak significant correlation coefficients (Pearson) were found between ΔDHEAS and ΔDunedinPACE for men (r=-0.18, p<0.001) and women (r=-0.11, p=0.018, Supplementary Table 2 and Supplementary Figure 1). After covariate adjustment ΔDHEAS showed an inverse association with ΔDunedinPACE in linear regression analyses for both men (β=-0.00002, p=0.022, Table 4) and women (β=-0.00003, p=0.005). Each one SD increase in DHEAS concentration over the assessment period was associated with a decrease in the pace of aging of 0.009 (men) and 0.01 (women) years, respectively, as measured by DunedinPACE. No further noteworthy associations were found between ΔDNAmAA and Δsex hormone levels (see Supplementary Table 5). Additionally, we conducted a subgroup analysis of participants who experienced a longitudinal hormone decline greater than the median (Supplementary Table 6). Here, no substantial differences from the overall findings were observed.

**Table 4:** Longitudinal multiple linear regression analyses of ΔDNAmAA on Δestradiol, ΔDHEAS and ΔFAI for older participants of the BASE-II.

| ΔHormone | ΔDNAmAA | Model | Estimate | SE | Men | | p-value | n | Estimate | SE | Women | | p-value | n |
| --- | --- | --- | --- | --- | --- | --- | --- | --- | --- | --- | --- | --- | --- | --- |
|  |  |  |  |  | CI<br>LL | CI<br>UL |  |  |  |  | CI<br>LL | CI<br>UL |  |  |
| Estradiol | Horvath | 1 | -3.8E-4 | 0.004 | -0.007 | 0.007 | 0.916 | 433 | -0.001 | 0.004 | -0.009 | 0.008 | 0.903 | 395 |
|  |  | 2 | -0.001 | 0.004 | -0.009 | 0.007 | 0.836 | 408 | -0.002 | 0.004 | -0.011 | 0.007 | 0.686 | 375 |
|  | GrimAge | 1 | -0.004 | 0.003 | -0.009 | 0.001 | 0.110 | 433 | -0.001 | 0.003 | -0.007 | 0.005 | 0.741 | 395 |
|  |  | 2 | -0.002 | 0.003 | -0.009 | 0.004 | 0.522 | 408 | 4.9E-4 | 0.004 | -0.006 | 0.007 | 0.890 | 375 |
|  | DunedinPACE | 1 | 1.7E-5 | 8.2E-5 | -1.4E-4 | 1.8E-4 | 0.832 | 472 | -8.3E-5 | 9.2E-5 | -2.6E-4 | 9.9E-5 | 0.371 | 455 |
|  |  | 2 | 9.5E-5 | 1.0E-4 | -1.1E-4 | 3.0E-4 | 0.357 | 445 | 5.3E-5 | 1.1E-4 | -1.6E-4 | 2.6E-4 | 0.620 | 424 |
| DHEAS | Horvath | 1 | 9.1E-5 | 3.9E-4 | -0.001 | 0.001 | 0.818 | 436 | -8.5E-5 | 0.001 | -0.001 | 0.001 | 0.874 | 394 |
|  |  | 2 | 2.6E-5 | 4.1E-4 | -0.001 | 0.001 | 0.950 | 412 | -1.6E-4 | 0.001 | -0.001 | 0.001 | 0.757 | 374 |
|  | GrimAge | 1 | -6.6E-5 | 2.9E-4 | -0.001 | 0.001 | 0.822 | 436 | -2.4E-4 | 3.9E-4 | -0.001 | 0.001 | 0.542 | 394 |
|  |  | 2 | -1.7E-4 | 3.2E-4 | -0.001 | 4.6E-4 | 0.604 | 412 | -3.8E-4 | 4.2E-4 | -0.001 | 4.4E-4 | 0.361 | 374 |
|  | DunedinPACE | 1 | -3.4E-5 | 8.7E-6 | -5.1E-5 | -1.7E-5 | <0.001 | 475 | -2.7E-5 | 1.1E-5 | -5.0E-5 | -4.6E-6 | 0.018 | 455 |
|  |  | 2 | -2.2E-5 | 9.6E-6 | -4.1E-5 | -3.2E-6 | 0.022 | 449 | -3.4E-5 | 1.2E-5 | -5.8E-5 | -1.0E-5 | 0.005 | 424 |
| FAI | Horvath | 1 | -0.003 | 0.012 | -0.027 | 0.021 | 0.794 | 430 | -0.134 | 0.120 | -0.370 | 0.101 | 0.263 | 377 |
|  |  | 2 | -0.009 | 0.017 | -0.042 | 0.024 | 0.605 | 405 | -0.069 | 0.219 | -0.499 | 0.361 | 0.752 | 359 |
|  | GrimAge | 1 | -0.008 | 0.009 | -0.026 | 0.010 | 0.374 | 430 | -0.007 | 0.087 | -0.179 | 0.164 | 0.932 | 377 |
|  |  | 2 | -0.012 | 0.013 | -0.038 | 0.014 | 0.359 | 405 | 0.109 | 0.171 | -0.227 | 0.445 | 0.522 | 359 |
|  | DunedinPACE | 1 | -3.6E-4 | 2.8E-4 | -0.001 | 1.8E-4 | 0.189 | 468 | 0.003 | 0.003 | -0.002 | 0.008 | 0.243 | 435 |
|  |  | 2 | -2.6E-4 | 4.0E-4 | -0.001 | 0.001 | 0.523 | 441 | 0.009 | 0.005 | -0.001 | 0.019 | 0.071 | 407 |

Model 1: unadjusted; Model 2: adjusted for chronological age at baseline, years since baseline assessment, baseline hormone concentration, baseline
DNAmAA, baseline smoking (pack years), alcohol (g/d), BMI. Note: DNAmAA=DNA methylation age acceleration, SE=Standard Error,
CI=Confidence Interval, LL=Lower Limit, UL=Upper Limit.

## Discussion

In this study, we investigated the relationship between estradiol, DHEAS and the FAI with biological age estimated from first-, second- and third-generation epigenetic clocks in cross-sectional and longitudinal data of BASE-II participants aged ≥60 years. Generally, no consistent and clinically relevant relationship was found between any of the sex hormones and DNAmAA estimators analysed in this study. Despite this overall lack of significant relationships we observed several nominal associations of unclear relevance. For instance, we identified a weak inverse association between DHEAS and Horvath’s DNAmAA in men at baseline. For women we found weak inverse associations between estradiol and DunedinPACE (baseline) and positive associations with GrimAge (follow-up). In longitudinal analyses, there was a weak inverse association between ΔDHEAS and ΔDunedinPACE, which was present in both sexes.

To date, only one study has assessed the relationship between epigenetic clocks as measures of biological aging and sex hormones. In that study by Kusters and colleagues, data of three population-based cohorts of similar age compared to the group of older BASE-II participants were examined cross-sectionally ^34^. They found that higher testosterone was associated with lower GrimAge and PhenoAge DNAmAA in men. For six more age acceleration measures they did not find significant associations ^34^. Regarding the age-related reduction in testosterone in men, studies found that lower testosterone levels were associated with increased mortality risk in older age ^23-25,42^. Testosterone replacement in testosterone deficient men, on the other hand, decreased mortality ^43,44^. Supporting this direction of effect, Kusters and colleagues suggested that the adverse health outcomes associated with lower testosterone might be moderated by accelerated epigenetic aging ^34^. While, in our fully adjusted models, we could not replicate this finding, the direction of the (non-significant) relationship of the FAI with GrimAge and DunedinPACE was in line with the results reported by Kusters and colleagues.

In our study, we observed an inverse association between estradiol and DunedinPACE in women at baseline, whereas at follow-up, estradiol levels were positively associated with the GrimAge clock. Given the inconsistent associations observed across our analyses and the small effect sizes, we consider it unlikely that these findings have relevant clinical implications. Furthermore, the lack of a meaningful relationship between estradiol and the DNAmAA estimators investigated here suggests that estradiol may not be a major driver of the accelerated rate of biological aging in women after the menopause ^33^. Regarding higher postmenopausal estradiol levels and the use of HRT, both health benefits and risks have been reported ^45-47^. Studies also suggested that survival benefits of postmenopausal HRT may diminish over time^48^. Thus, while estradiol’s influence on aging is complex, our results do not support a strong role for estradiol in mediating these effects through the DNAmAA mechanisms in the older women studied here.

The reputation of DHEAS as an anti-aging hormone stems from studies that have found a higher mortality risk ^19,49,50^ and a reduced physical capacity in association with lower DHEAS levels ^19,51^. Fahy and colleagues published a study on epigenetic age reversal involving DHEA supplementation suggesting a potential positive impact of higher DHEAS levels on biological aging ^52^. Therefore, we assumed an inverse association to be present in our data. Indeed, we found higher DHEAS levels associated with slower epigenetic aging as measured by Horvath’s clock in cross-sectional analyses in men. With each one SD increase in DHEAS, Horvath’s DNAmAA decreased by 0.45 years. Also, participants of both sexes exhibiting an increase in DHEAS concentration of one SD over the assessment period of on average 7.3 years, decreased in their pace of aging (DunedinPACE) by 0.009 and 0.01 years, respectively. Our findings suggest that increases in DHEAS levels are associated with a slightly slower rate of biological aging (DunedinPACE and Horvath’s DNAmAA) thereby supporting the direction of an effect reported before. However, the effect size estimate was relatively small, and additional research would be needed to further investigate clinical relevance and potential underlying mechanisms.

We note that the lack of consistent associations in this study does not necessarily exclude the presence of a relationship between sex hormone levels and DNAmAA. The lack of noteworthy findings may also be due to one or several of the limitations of our study. First, we were not able to investigate bioavailable hormones due to lack of albumin measurement at follow-up. However, as a proxy and commonly used parameter to estimate the unbound testosterone fraction we used the FAI in our study ^35^. Second, around a third of the baseline participants were not followed up, which possibly introduces a selection bias. However, a comparison of sex hormone levels between participants with baseline and follow-up data and those lost to follow-up revealed no statistically significant differences, except that the dropout group had a higher average age (Supplementary Table 7). Third, despite the fact that with n=1,404 our sample size was relatively large, we cannot exclude the possibility that the lack of significant findings may be due to insufficient power, especially to detect small effects. Finally, as previously reported, the study population investigated here tends to have an above-average health status, which limits the generalisability of the results to the general population.

Strengths of this study include the availability of six different epigenetic clocks and four sex hormones at two time points with a mean follow-up time of 7.3 years. This allowed us to investigate the long-term relationships between sex hormones and DNAmAA, in addition to cross-sectional analyses, which was not done before. Moreover, due to the detailed information on hormone intake and medication, it was possible to exclude participants who were taking exogenous sex hormones. Information on potentially confounding factors such as BMI, cigarette smoking and alcohol intake were available, which allowed us to control for these factors.

The age-related decline in sex hormones has substantial implications for the health and quality of life in the older population. Our study provides explorative insights into the cross-sectional and longitudinal relationship between sex hormones and DNAmAA in older adults. Future studies with longitudinal measurements of sex hormones and DNAmAA in a study population that includes participants at the age of hormonal transition are needed to further investigate the relationship between biological aging and sex hormone levels.

## Conclusion

In this study, we cross-sectionally and longitudinally investigated the relationship between epigenetic clocks and sex hormones in 1,404 participants aged ≥60 years over a 7.3-year follow-up period. Generally, we did not observe a consistent relationship between estradiol, DHEAS, the FAI and epigenetic age as assessed by six different DNAmAA estimators. This suggests that sex hormones play at best a minor role with respect to biological aging in the older population studied here.

## Data Availability

All data produced in the present study are available upon reasonable request to the authors.

## Acknowledgments

This work was supported by grants of the Deutsche Forschungsgemeinschaft (project number 460683900 to ID & LB), the ERC (as part of the Lifebrain project to LB), and the Cure Alzheimer’s Fund (as part of the CIRCUITS consortium to LB). Additional funds supporting this research came from a grant of the EU Joint Programme – Neurodegenerative Disease Research (project EPIC4ND, coordination: C.M.L.). This article uses data from the Berlin Aging Study II (BASE-II) and the GendAge study, which were supported by the German Federal Ministry of Education and Research under grant numbers #01UW0808; #16SV5536K, #16SV5537, #16SV5538, #16SV5837, #01GL1716A and #01GL1716B. We thank all probands of the BASE-II/GendAge study for their participation in this research. C. M. L. was supported by the Heisenberg programme of the German Research Foundation (DFG; LI 2654/4-1).

## Author contributions

Conceived and designed the study: HS, VMV and ID. Contributed study-specific data: VMV, JH, VRZ, LB, and ID. Analysed the data: HS. Wrote the manuscript: HS, VMV and ID. All authors revised and approved the manuscript.

## Data availability statement

Due to concerns for participant privacy, data are available only upon reasonable request. Please contact Ludmila Müller, scientific coordinator, at, for additional information.

## Ethics

All participants gave written informed consent. Medical assessments were conducted in accordance with the Declaration of Helsinki and approved by the Ethics Committee of the Charité – Universitaetsmedizin Berlin (approval numbers EA2/029/09 and EA2/144/16) and were registered in the German Clinical Trials Registry as DRKS00009277 and DRKS00016157.

