## Supplemental Methods for "Cross-sectional and Longitudinal Relationship between Sex Hormones and Six Epigenetic Clocks in Older Adults: Results of the Berlin Aging Study II (BASE-II)"

**Corresponding author:**

| | Hormone | Method | Detection range |
|---|---|---|---|
| Until September 2013 | Estradiol | Fluoroimmunoassay, AutoDELFIA system (PerkinElmer Inc., Waltham, MA)<br>Cat. No. B056-101 | 14.98 to 1446.44 pg/ml |
| Until September 2013 | Total Testosterone | Fluoroimmunoassay, AutoDELFIA, system (Perkin Elmer Inc., Waltham, MA)<br>Cat. No. B050-201 | 0.1296 to 17.91 ng/ml, concentrations below the detection limit defined as 0.05 ng/ml |
| Until February 2014 | DHEAS | Radioimmunoassay, TKDS Coat-A-Count kits (Siemens Healthcare Diagnostics Ltd., Germany)<br>Cat. No. TKDS1 | 5 to 1000 µg/dl |
| Until September 2013 | SHBG | Chemiluminescent immunoassay, Immulite 2000 system (Siemens Healthcare Diagnostics Products Ltd., Germany)<br>Cat. No. L2KSH2 | 0.02 to 180 nmol/l |
| New method | Estradiol | Electrochemiluminescence immunoassays (ECLIA), Elecsys and cobas e (Roche Diagnostics GmbH, Mannheim, Germany)<br>Ref. 03000079 122 | 18.4 to 11010 pmol/l, concentrations below the detection limit defined as 9.2 pmol/l |
| New method | Total Testosterone | Electrochemiluminescence immunoassays (ECLIA), Elecsys and cobas e (Roche Diagnostics GmbH, Mannheim, Germany)<br>Ref. 05200067 190 | 0.025 to 15 ng/ml, concentrations below the detection limit defined as 0.015 ng/ml |
| New method | DHEAS | Electrochemiluminescence immunoassays (ECLIA), Elecsys and cobas e (Roche Diagnostics GmbH, Mannheim, Germany)<br>Ref. 03000087 122 | 4 to 10000 ng/ml |

| | Hormone | Method | Detection range |
| --- | --- | --- | --- |
| New method | SHBG | Electrochemiluminescence immunoassays (ECLIA), Elecsys and cobas e (Roche Diagnostics GmbH, Mannheim, Germany)<br>Ref. 03052001 190 | 0.35 to 200 nmol/l |

Note. All measurements were performed in serum samples
